# Multimodal Immune Profile Score (IPS) as a Pan-Cancer Biomarker for Stratifying Outcomes of Immune Checkpoint Inhibitor Benefit in Microsatellite Stable Colorectal Cancer and Rare Solid Tumors

**DOI:** 10.64898/2026.10.07.26364989

**Authors:** Michelle A. Ting-Lin, Emmanuel S. Antonarakis, Xifeng Wang, Rossin Erbe, Alia D. Zander, Yan Liu, Ailin Jin, Gregory A. Vidal, Scott M. Haake, Michelle M. Stein, Dana F. DeSantis, Kyle A. Beauchamp, Matthew E. Campbell, Ezra E.W. Cohen, Seung Won Hyun, Ronan J. Kelly, Aparna Kalyan, Babar Bashir, B.J. Rimel, Thomas J. Herzog, Victoria L. Chiou, Chithra Sangli, Halla Nimeiri, Jay Carlson, Diane M. Simeone, John H. Strickler

**Affiliations:** Tempus AI, Inc.; University of Minnesota, Minneapolis, MN; Clinical Research and Breast Oncology Program, Lee S Schwartzberg Research Center, West Cancer Center and Research Institute, Memphis, TN; Vanderbilt Health, Nashville, TN, USA; Nashville VA Hospital, Nashville, TN; Charles A. Sammons Cancer Center, Baylor University Medical Center, Dallas, Texas; Northwestern University Feinberg School of Medicine, Robert H. Lurie Comprehensive Cancer Center, Chicago, IL; Thomas Jefferson University, Philadelphia, PA; University of Washington and Fred Hutch Cancer Center, Seattle, WA; University of Cincinnati Cancer Center; University of Cincinnati, Cincinnati, OH; Mercy Health, St. Louis, MO; UCSD Moores Cancer Center, San Diego, CA; Duke University, Durham, NC

## Abstract

**Background:** Immune checkpoint inhibitor (ICI) therapies have revolutionized oncology, but identifying patients who benefit remains a challenge, particularly in populations where ICIs are not the standard of care, including microsatellite stable colorectal cancer (MSS CRC) and rare solid cancers. We evaluated real-world performance of the Immune Profile Score (IPS), a validated DNA- and RNA-based molecular signature, in these cohorts.

**Methods:** In an exploratory analysis, we analyzed two real-world cohorts: 1) MSS CRC; and 2) MSS, tumor mutational burden-low rare solid cancers treated with off-label ICI. IPS-High and IPS-Low were calculated using a previously validated threshold. Cox proportional hazards models were fit to demonstrate prognostic utility for real-world overall survival (rwOS). In MSS CRC, time-to-next-treatment (TTNT) on prior non-ICI therapy was compared to rwOS on subsequent ICI therapy.

**Results:** In MSS CRC (n=46), IPS-High patients (13%) demonstrated clinically meaningful improvement in rwOS versus IPS-Low (Median OS: not reached vs 7.3 months; HR = 0.22, 90% CI 0.04-1.16). While IPS was not associated with TTNT on the preceding non-ICI line (HR = 1.07, 90% CI 0.60-1.91), it was associated with OS in patients receiving later line ICI (HR 0.21, 90% CI 0.04-1.22). In the rare cancers cohort (n=90; 26 unique malignancies), IPS-High (17.8%) was associated with longer OS to ICI therapy (HR = 0.26, 90% CI 0.11-0.61), even when restricted to histologies represented in both IPS groups (HR = 0.18, 90%, CI 0.05-0.62).

**Conclusions:** Preliminary evidence suggests that IPS may serve as a generalizable, pan-cancer biomarker capable of stratifying outcomes for ICI treatment in populations traditionally excluded from immunotherapy benefit.

## Background

The clinical use of immune checkpoint inhibitors (ICIs) has significantly advanced oncology; however, substantial challenges persist in identifying patients most likely to benefit, particularly in cohorts where ICIs are not yet the standard of care. Conventional biomarkers, like tumor mutational burden (TMB) and PD-L1, often fail to capture the complexity of the tumor microenvironment. High-throughput multimodal data have enabled a comprehensive understanding of patients. Integrating these multimodal data, encompassing molecular, histopathology, radiology, and clinical records, with deep learning methodologies can help elucidate the biological processes implicated in multifactorial diseases such as cancer [1].

We previously described the development and validation of a multiomic, pan-solid cancer biomarker, the Immune Profile Score (IPS) algorithm. The IPS model includes TMB and 11 RNA-based biomarkers as features; IPS is calculated as a linear predictor by summing these 12 features weighted by their respective Cox coefficients and is then scaled to range from 0 and 100. In the preceding clinical validation study, IPS was established as a robust, DNA- and RNA-based molecular signature capable of predicting survival outcomes across pan-cancer solid tumors even beyond current standard of care biomarkers [2].

Here we further tested the real-world performance of IPS in two cohorts with a high degree of clinical unmet need: patients with microsatellite stable colorectal cancer (MSS CRC) and patients with MSS, TMB-low rare solid cancers. The efficacy of ICIs in MSS CRC remains disappointingly low, with objective response rates to ICI monotherapy typically below 10% [3], and, the majority of CRC patients (approximately 85%) have MSS tumors proficient in mismatch repair, which is associated with significantly poorer prognoses compared to MSI-H CRC [4,5]. This stark contrast in prognosis underscores a critical need for robust biomarkers to predict immunotherapy response in MSS CRC.

Similarly, rare cancers, defined by the FDA in the Orphan Drug Act as affecting fewer than 200,000 global cases annually, frequently lack extensive clinical trial data to guide immunotherapy utilization. While landmark efforts like the SWOG S1609 DART trial successfully evaluated efficacy signals across 53 refractory rare cancer cohorts treated with dual CTLA-4 and PD-1 inhibition, the heterogeneous outcomes observed across histologies underscore the need for predictive biomarkers to refine patient selection [6]. Rare cancers collectively account for approximately 25% of all adult oncology diagnoses and are associated with a significantly lower 5-year survival rate compared to more common malignancies [7]; more effective therapeutic strategies are critically needed.

Leveraging the Tempus clinicogenomic database, we evaluated whether IPS-high (IPS-H) status correlates with improved immunotherapy outcomes in these two distinct patient cohorts.

## Methods

### Cohort Selections

Two cohorts (**Figure 1**) representing conditions with high unmet clinical need and for which ICI therapy is not approved were identified from the Tempus clinicogenomic database after applying clinically relevant inclusion criteria: cohort 1) microsatellite stable colorectal cancer (MSS CRC); and cohort 2) MSS and TMB-low (TMB-L) rare solid cancer.

**Figure 1.**
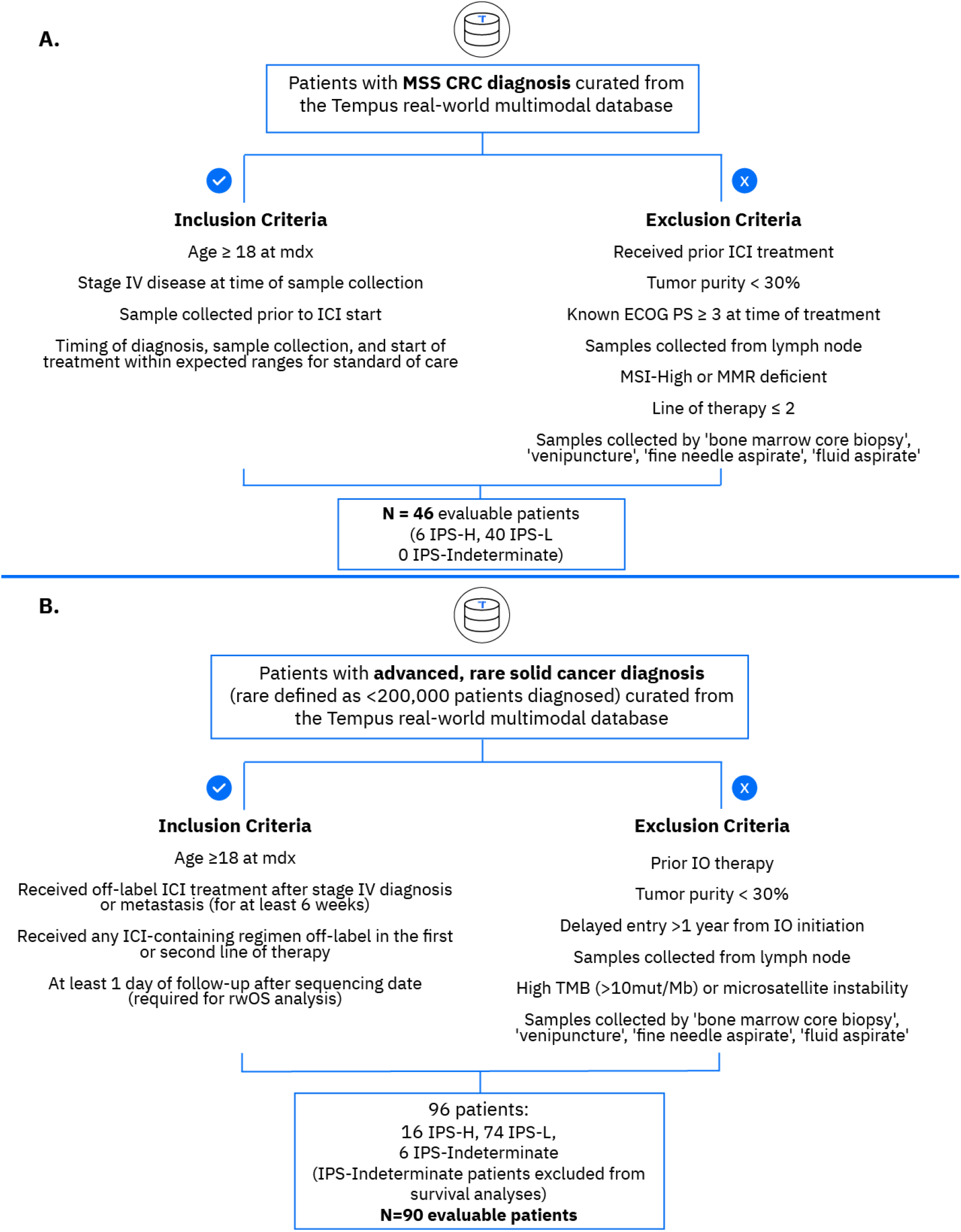
Cohort funnel. (a) MSS CRC cohort funnel. (b) Rare cancer cohort funnel.

Patients in cohort 1 were diagnosed with stage IV MSS CRC and received ICI for a minimum of 2 cycles or 6 weeks in the third-line (3L) setting or beyond. Patients were limited to the 3L setting and beyond to specifically mimic the refractory population typical of late-phase clinical trials in MSS CRC. Patients in cohort 2 were diagnosed with stage IV cancer, defined as rare per the FDA (fewer than 200,000 cases annually), and received off-label ICI for a minimum of 2 cycles or 6 weeks in the 1L or 2L setting to minimize confounding influence of multiple prior systemic therapies. Patients exhibiting high TMB (>10 mut/Mb) or microsatellite instability high status were excluded from cohort 2 due to the existing histology agnostic biomarker-based approvals. The specific cancer types included in cohort 2 are listed in **Supplemental Table 1**. Exclusion criteria for both cohorts included low tumor purity (<30%) and samples derived from cytology or lymph node biopsies due to ambiguity of anatomic location of lymph node biopsy and high expression of immune genes in the lymph node (**Figure 1**).

### IPS Calculation

IPS is a validated, machine learning developed, DNA- and RNA-based molecular signature that has been described in detail previously [2]. Briefly, IPS was calculated as a score on a continuous scale from 0-100 where a score of 0-44 was IPS-Low (IPS-L), 48-100 was IPS-High (IPS-H), and 45-47 was indeterminate. The indeterminate zone is a locked analytical parameter that serves to minimize changes in IPS score between IPS-L and IPS-H when accounting for small variability that may be observed when sequencing the same sample multiple times. IPS model features include markers of immune inflammation (e.g. *CD274*), T-cell exhaustion (e.g. *CD276, IDO1*), neoantigen burden (TMB), and tumor immune resistance (“immune exhaustion signature”).

### Statistical Analyses

Eligibility criteria and outcomes analyses were pre-specified to aid in future prospective trial planning if results were promising. Descriptive statistics were used to summarize clinical characteristics for both cohorts.

#### Cohort 1

A Cox proportional hazards model was utilized to compare overall survival (OS) between IPS-H and IPS-L patients, with IPS serving as the sole binary independent variable. OS was defined as the time from the index date (the initiation date of ICI) to the date of death from any cause. The study period was defined to be 24 months from initiation of ICI. For patients whose sequencing occurred after the initiation of ICI, risk set adjustment was applied to account for the delayed entry. In an exploratory analysis of predictive utility, the hazard ratio (HR) for OS during ICI treatment was compared to the HR for time to next treatment (TTNT) during the preceding line of non-ICI therapy. TTNT was defined as the time from prior non-ICI initiation date to the ICI initiation date and used as a surrogate for progression-free survival.

#### Cohort 2

A stratified Cox proportional hazards model was employed for the OS analysis, with adjustment for treatment regimen (ICI monotherapy vs. ICI combination) and stratification by line of therapy (1L vs. 2L). The OS definition and delayed entry adjustments were applied identically to the MSS CRC cohort. Additionally, two separate OS analyses were conducted: one evaluating the binary IPS (IPS-H vs. IPS-L) within a restricted subgroup of cancer types that included patients from both IPS status, and another utilizing the continuous IPS score in place of the binary IPS categorization across the overall cohort. Six patients were IPS-indeterminate, and subsequently excluded from further analyses comparing IPS-H to IPS-L.

This study represents an exploratory, retrospective analysis aimed at hypothesis generation; formal statistical hypothesis testing was not performed. Consequently, all reported survival estimates and confidence intervals should be interpreted as exploratory.

### Data Availability and Ethics Statement

De-identified data used in this research were collected in a real-world healthcare setting and subject to controlled access for privacy and proprietary reasons. When possible, derived data supporting the findings of this study have been made available within the paper and its online supplemental figures and tables. This study was conducted in accordance with HIPAA regulations where applicable, and an IRB-exempt determination (Advarra Pro00072742).

## Results

### Patient characterization of cohorts

The MSS CRC cohort comprised 46 ICI-treated patients with a median follow up of 8.1 months: median (IQR) age of 54 (39.0-83.0), 48% female (n=22), 26 (57%) were white, and 38 (83%) had liver metastasis (**Table 1**). Six (13%) patients were IPS-H. TMB data were available on all patients in the cohort, and a higher proportion of IPS-H patients were TMB-L (n=4 (67%)) versus TMB-H (n=2 (33%)) (**Table 1**). Lines of therapy were well balanced between 3L (n=16 (34.8%)), 4L (n=15 (32.6%)) and 4L+ (n=15 (32.6%)).

**Table 1:** Cohort clinical characteristics.

| Table 1. Cohort Clinical Characteristics |  |  |  |  |  |  |  |
| --- | --- | --- | --- | --- | --- | --- | --- |
|  | MSS Cohort |  |  | Rare Cancers Cohort |  |  |  |
|  | Overall<br>N = 46 | IPS-High<br>n = 6 | IPS-Low<br>n = 40 | Overall<br>N = 96 | IPS-High<br>n = 16 | IPS-Low<br>n = 74 | IPS-Indeterminate<br>n = 6 |
| **Age** |  |  |  |  |  |  |  |
| Median<br>(Min-Max) | 54.0<br>(39.0-83.0) | 49.0<br>(43.0-73.0) | 54.5<br>(39.0-83.0) | 64.5<br>(27.0-84.0) | 66.5<br>(37.0-81.0<br>) | 63.5<br>(27.0-84.<br>0) | 66.0<br>(39.0-77.0) |
| **Sex** |  |  |  |  |  |  |  |
| Male | 24 (52.2%) | 2 (33.3%) | 22 (55.0%) | 30 (31%) | 6 (38%) | 23 (31%) | 1 (17%) |
| Female | 22 (47.8%) | 4 (66.7%) | 18 (45.0%) | 66 (69%) | 10 (62%) | 51 (69%) | 5 (83%) |
| **Race** |  |  |  |  |  |  |  |
| White | 26 (56.5%) | 2 (33.3%) | 24 (60.0%) | 45 (47%) | 7 (44%) | 35 (47%) | 3 (50%) |
| Black or African<br>American | 4 (8.70%) | 1 (16.7%) | 3 (7.50%) | 10 (10%) | 0 (0%) | 10 (14%) | 0 (0%) |
| Asian | 5 (10.9%) | 1 (16.7%) | 4 (10.0%) | 1 (1.0%) | 1 (6.3%) | 0 (0%) | 0 (0%) |
| Other Race | 1 (2.17%) | 0 (0%) | 1 (2.50%) | 12 (13%) | 5 (31%) | 7 (9.5%) | 0 (0%) |
| Unknown/<br>missing | 10 (21.7%) | 2 (33.3%) | 8 (20.0%) | 28 (29%) | 3 (19%) | 22 (30%) | 3 (50%) |
| **ECOG Score** |  |  |  |  |  |  |  |
| 0 | 10 (21.7%) | 3 (50.0%) | 7 (17.5%) | 32 (33%) | 3 (19%) | 28 (38%) | 1 (17%) |
| 1 | 18 (39.1%) | 3 (50.0%) | 15 (37.5%) | 23 (24%) | 7 (44%) | 14 (19%) | 2 (33%) |
| 2 | 4 (8.70%) | 0 (0%) | 4 (10.0%) | 4 (4.2%) | 1 (6.3%) | 3 (4.1%) | 0 (0%) |
| Unknown | 14 (30.4%) | 0 (0%) | 14 (35.0%) | 37 (39%) | 5 (31%) | 29 (39%) | 3 (50%) |
| **LOT** |  |  |  |  |  |  |  |
| 3 (MSS) \| 1 (rare) | 16 (34.8%) | 0 (0%) | 16 (40.0%) | 62 (65%) | 12 (75%) | 47 (64%) | 3 (50%) |
| 4 (MSS) \| 2 (rare) | 15 (32.6%) | 2 (33.3%) | 13 (32.5%) | 34 (35%) | 4 (25%) | 27 (36%) | 3 (50%) |
| 4L+ (MSS) \| x (rare) | 15 (32.6%) | 4 (66.7%) | 11 (27.5%) | x | x | x | x |
| **Liver Metastasis**<br>**Documented** |  |  |  |  |  |  |  |
| Yes | 38 (82.6%) | 3 (50.0%) | 35 (87.5%) | 40 (42%) | 3 (19%) | 34 (46%) | 3 (50%) |
| No | 8 (17.4%) | 3 (50.0%) | 5 (12.5%) | 56 (58%) | 13 (81%) | 40 (54%) | 3 (50%) |
| **Lung Metastasis**<br>**Documented** |  |  |  |  |  |  |  |
| **Yes** | 27 (59%) | 5 (83%) | 22 (55%) | 37 (39%) | 9 (56%) | 25 (34%) | 3 (50%) |
| **No** | 19 (41%) | 1 (17%) | 18 (45%) | 59 (61%) | 7 (44%) | 49 (66%) | 3 (50%) |
| **Therapy Type** |  |  |  |  |  |  |  |
| ICI + Additional | 22 (47.8%) | 3 (50.0%) | 19 (47.5%) | 72 (75%) | 10 (63%) | 57 (77%) | 5 (83%) |
| ICI-Only | 24 (52.2%) | 3 (50.0%) | 21 (52.5%) | 24 (25%) | 6 (38%) | 17 (23%) | 1 (17%) |
| **ICI specific class** |  |  |  |  |  |  |  |
| PD1 | 41 (89%) | 6 (100%) | 35 (88%) | 62 (65%) | 13 (81%) | 45 (61%) | 4 (67%) |
| PDL1 | 2 (4%) | 0 (0%) | 2 (5%) | 26 (27%) | 3 (19%) | 22 (30%) | 1 (17%) |
| CTLA4+PD1 | 2 (4%) | 0 (0%) | 2 (5%) | 7 (7%) | 0 (0%) | 7 (9%) | 0 (0%) |
| Other | 1 (2%) | 0 (0%) | 1 (3%) | 1 (1%) | 0 (0%) | 0 (0%) | 1 (17%) |
| **TMB** |  |  |  |  |  |  |  |
| High | 8 (17.4%) | 2 (33.3%) | 6 (15.0%) | x | x | x | x |
| Low | 38 (82.6%) | 4 (66.7%) | 34 (85.0%) | x | x | x | x |

The rare cancers cohort comprised 96 ICI-treated patients with a median follow up of 20 months: median (IQR) age of 64 (55.8-71.0), 69% female (n=66), 45 (47%) were white, and 40 (42%) had liver metastasis (**Table 1**). There were 26 cancer types included in the cohort, with notable representation of gynecologic malignancies, which collectively accounted for 36% (n=35) of the patients (**Supplemental Table 1)**. The most common cancer was carcinosarcoma (n=19 (20%)) followed by pancreatic adenocarcinoma (n=17 (18%)). 16/90 (17.8%) patients were classified as IPS-H. Only 7 cancer types had representation from both IPS-H and IPS-L (**Supplemental Table 2)**. Of the ICI regimen types, ICI+CT (n=53 (55%)) was the most common, followed by ICI monotherapy (n=19 (20%)) and ICI+TKI (n=17 (18%)). The majority of patients (n=62 (65%)) received ICI regimen in 1L. The majority of patients (n=29 (85%)) who received ICI regimen in the 2L were treated with chemotherapy in 1L (**Table 1**).

### Overall survival

In the MSS CRC cohort, a clinically meaningful improvement in real-world survival was observed in the IPS-H versus IPS-L groups (**Figure 2 a**). The median overall survival (OS) was not reached within a 24-month follow-up period for the IPS-H group, compared to 7.7 months for the IPS-L group. This effect size (HR=0.22, 90% CI 0.04-1.16) was comparable to our prior validation of IPS stratification across advanced solid cancers (HR=0.45, 90% CI 0.40-0.52) and remained consistent when adjusting for therapy line and therapy type (HR=0.20, 90% CI 0.04-1.09).

**Figure 2.**
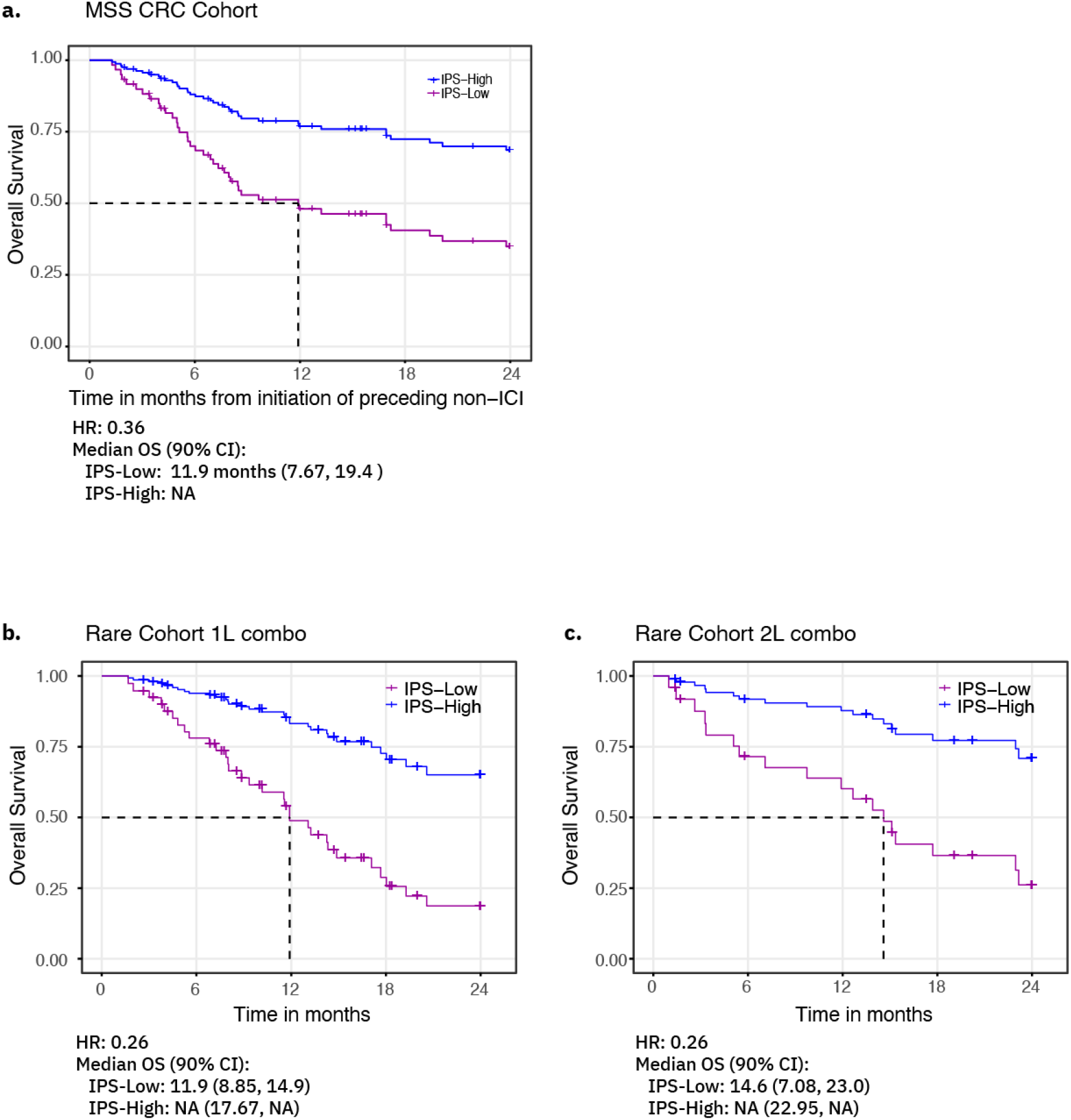
OS KM curves between IPS-H and IPS-L. **Overall Survival** (**a**) IPS-H MSS CRC patients have longer OS compared to IPS-low. Cox Proportional Hazards model curves for real world overall survival stratification of IPS fit to advanced/metastatic MSS CRC patients treated with ICI (N = 46). The Hazard Ratio is 0.22, 90% Confidence Interval (CI) (0.04, 1.16). Median OS for IPS-Low is 7.67, 90% CI (5.74, 13.2) months. Median OS for the IPS-High group was not reached in 24 months. IPS-H patients with rare cancers demonstrated clinically improved overall survival compared to IPS-L patients. Those who received 1L combo (**b**) and patients who received 2L combo therapy (**c**) shown as representative examples. Patients who received 1L and 2L monotherapy showed similar trends for predicted overall survival. Hazard ratios (HR) and median OS with 90% confidence intervals (CI) are shown on the graphs.

In the rare cancers cohort, a multivariate Cox Proportional Hazards (PH) model, controlling for regimen (ICI monotherapy or ICI combined with other therapies) and stratified by line of therapy (1L or 2L), was utilized to assess the prognostic association of the IPS-H and IPS-L classifications with OS. Among patients receiving ICI combination therapy in the first (**Figure 2 b**) or second line (**Figure 2 c**), OS was clinically improved in IPS-H patients compared to IPS-L.

Additionally, OS was clinically improved in patients whose tumors were classified as IPS-H compared to those classified as IPS-L (HR=0.26, 90% CI 0.11 to 0.61). The prognostic association of IPS persisted when restricting the analysis to cancer types with representation from both IPS-H and IPS-L (HR=0.18, 90% CI 0.05 to 0.62). Finally, we evaluated the prognostic utility of IPS as a continuous variable. The hazard ratio (for a 50-unit increase, HR=0.15, 90% CI 0.06 to 0.38) exhibited a similar magnitude and direction to the categorical representation comparing IPS-H versus IPS-L.

### Evaluation of predictive utility for IPS

In the MSS CRC cohort, to test the potential predictive utility of IPS, we compared IPS risk stratification on ICI therapy to the prior non-ICI regimen. While IPS was not associated with TTNT on non-ICI therapy in the preceding line (HR=1.07, 90% CI 0.60-1.91), it was associated with OS in patients on ICI therapy (rwOS HR = 0.21, 90% CI 0.04-1.22) (**Figure 3)**. The difference in stratification between the non-ICI prior treatment cohort versus the ICI cohort was clinically meaningful. The analysis of predictive utility, comparing outcomes on ICI therapy to a preceding line of non-ICI therapy, was conducted only within the MSS CRC cohort. This analysis was not feasible for the rare cancer cohort, as the majority of these patients received ICI as their 1L therapy, precluding a baseline comparison against prior systemic treatment.

**Figure 3.**
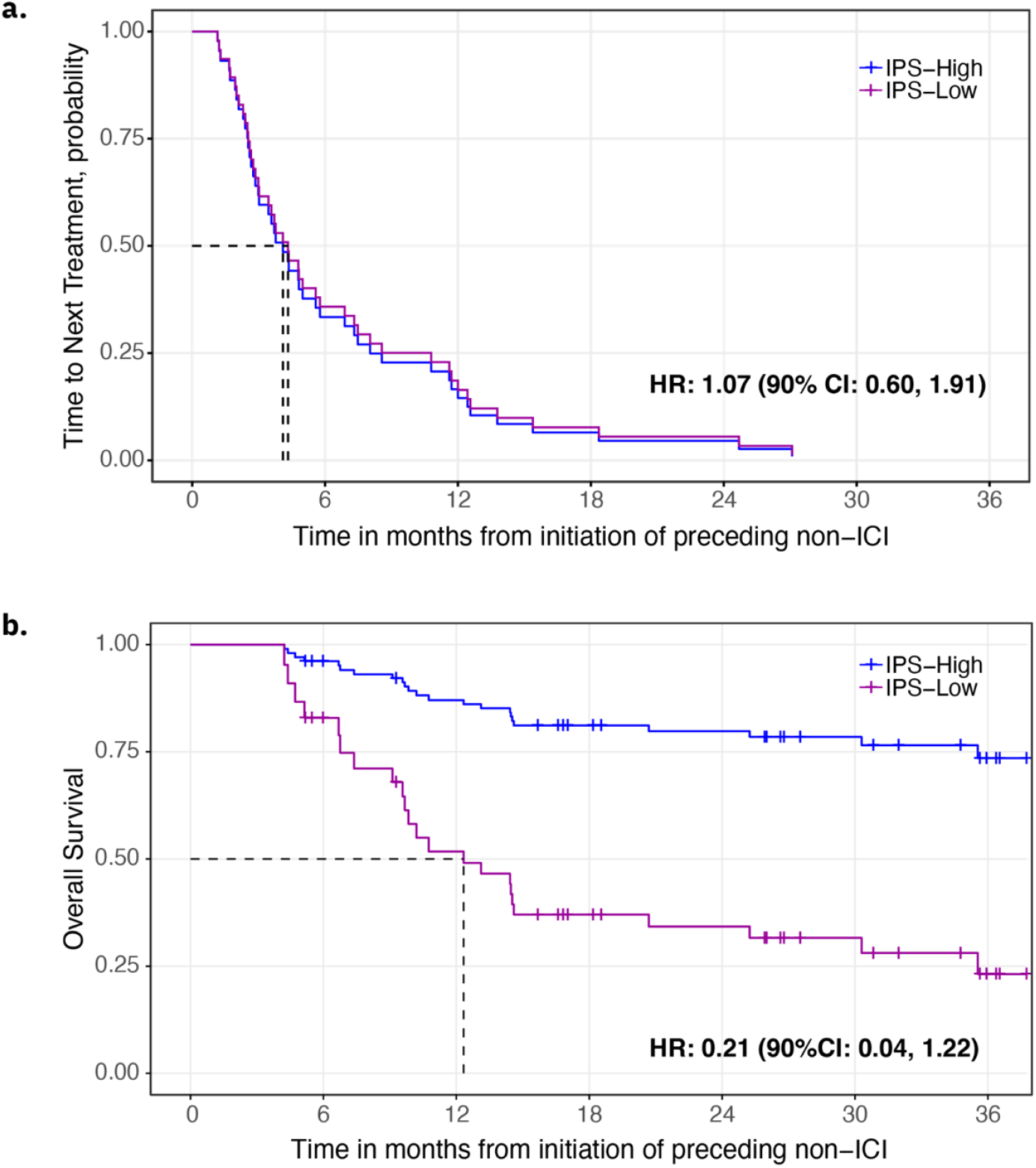
OS by IPS-H vs IPS-L on ICI compared to TTNT on prior non-ICI. **TTNT and OS**. Cox Proportional Hazards model curves stratified by IPS fit to (a) Real-world time to next treatment (TTNT) for MSS CRC patients on the treatment line prior to ICI therapy. The Hazard Ratio (90% CI) is 1.07 (0.60, 1.91). (b) Real-world overall survival for ICI treated MSS CRC patients. The Hazard Ratio (90% CI) is 0.21 (0.04, 1.22)

## Discussion

This study extends our initial clinical validation of the IPS algorithm by demonstrating its utility in populations traditionally excluded from deriving benefit from immunotherapy. By identifying patients most likely to respond to ICI in non-standard-of-care, IPS may facilitate label expansion of ICIs to include a subset of patients with MSS CRC and rare cancer types. These real-world data support the continued integration of multiomic scoring systems to guide personalized immunotherapy strategies in areas where more effective therapeutic options are critically needed.

IPS-H status effectively identified a distinct and clinically relevant subset of MSS CRC patients who may achieve a clinically meaningful survival benefit from ICI therapy. The separation in survival outcomes within the MSS CRC cohort (not reached for IPS-H vs. 7.67 months for IPS-L) challenges the assumption that MSS status precludes a meaningful response to ICIs in mCRC. The IPS-H MSS CRC patients exhibited survival curves mirroring those of MSI-H patients in literature [8–10]. Notably, despite the presence of liver metastasis in 83% of the total cohort and 50% of the IPS-H subset, IPS-H status remained a powerful predictor of survival [11]. Furthermore, our analysis demonstrates initial evidence for the predictive nature of the IPS biomarker. By comparing the outcomes of IPS-H and IPS-L patients while on ICI therapy to their preceding outcomes on non-ICI systemic treatments, we established that the benefit is specifically tied to the ICI mechanism of action. Longer survival observed in the IPS-H group suggests an enhanced responsiveness to immunotherapy, rather than better underlying overall health or less aggressive disease biology. Taken together, this could shift treatment paradigms in clinically advanced settings, offering a molecularly-guided alternative to standard salvage therapies for MSS-CRC patients who are found to be IPS-H.

The scarcity and biological heterogeneity of rare cancers present challenges to widespread therapeutic advancement, often leaving clinicians without clear evidence-based guidelines. A systematic review of ICI efficacy in rare tumors noted that a lack of biomarker-driven studies is a major limitation in the field [12]. Our MSS, TMB-L rare cancers cohort, encompassing 26 distinct malignancies, reinforces the robust, histology-agnostic performance of the IPS algorithm. The compelling performance of IPS across diverse histologies (HR=0.26) remained significant (HR=0.18) even when restricting the analysis to cancer types represented in both IPS-H and IPS-L groups. By identifying 17% of patients with advanced rare solid tumors as IPS-H, IPS captures a responder population that would otherwise be overlooked by conventional predictive biomarkers and histology-specific guidelines. Future prospective early-phase basket trials of ICI with chemotherapy could benefit from these data.

The MSS CRC analysis was based on 46 patients, of whom only six (13%) were classified as IPS-H. The small number of IPS-H patients resulted in relatively wide confidence intervals, suggesting that, while the effect size is clinically meaningful, the precision is limited by the sample size. To address this, we have initiated a prospective-retrospective validation using a randomized phase II clinical trial cohort with a larger, more uniform sample and a randomized control arm to definitively establish the predictive utility of IPS. While the rare cancer cohort was larger, it was highly heterogenous, encompassing 26 different cancer types; only seven cancer types had representation from both IPS-H and IPS-L patients, which limits the ability to draw definitive conclusions for specific histologies. Therefore, conclusions drawn from this cohort should be interpreted as generalized findings across a diverse group of rare malignancies, rather than as definitive proof for any particular single histology.

In summary, we demonstrated across two diverse real-world clinical datasets that IPS functions as a generalizable, pan-cancer biomarker capable of accurately stratifying outcomes for ICI treatment across various cancer types and cohorts beyond our original clinical validation study. To our knowledge we are the first to report a pan-cancer biomarker beyond TMB for ICI in rare cancers. Further prospective-retrospective validation studies are currently in progress to strengthen the clinical applicability of these real-world findings.

## Data Availability

De-identified data used in this research were collected in a real-world healthcare setting and subject to controlled access for privacy and proprietary reasons. When possible, derived data supporting the findings of this study have been made available within the paper and its online supplemental figures and tables.

## Author Contributions

MAT-L and DFD wrote the original draft of the manuscript. ADZ, RE, XW, YL, AJ, SWH, MMS, KB and CS performed research and analyses. MAT-L, ADZ, RE, YL, MMS, CS, HN, and EEWC participated in conception and design of the work. CS, VC, HN, and EEWC performed supervisory roles. ESA, SMH, BJR, TH, JC, JHS, GV, RJK, DS, AK, and BB provided clinical insights and critical commentary on the manuscript. All authors reviewed and approved the final version of this manuscript. HN (Tempus AI) is the guarantor for the manuscript.

## Acknowledgments

We thank Ben Terdich for operations support and members of the Tempus labs for sample sequencing. Parts of this work were presented at SITC 2025 and ASCO 2026. There is no funding associated with this work.

