## Supplemental Information for "Multimodal Immune Profile Score (IPS) as a Pan-Cancer Biomarker for Stratifying Outcomes of Immune Checkpoint Inhibitor Benefit in Microsatellite Stable Colorectal Cancer and Rare Solid Tumors"

### Supplemental Materials

**Supplemental Table 1. Cancers included in rare cancers cohort**

| Cancer Types | Overall N = 96 | IPS-High n = 16 | IPS-Low n = 74 | IPS-Indeterminate n = 6 |
|---|---|---|---|---|
| adenoid cystic carcinoma | 2 (2.1%) | 0 (0%) | 2 (2.7%) | 0 (0%) |
| adrenal cortical carcinoma | 3 (3.1%) | 0 (0%) | 3 (4.1%) | 0 (0%) |
| anaplastic thyroid cancer | 1 (1.0%) | 0 (0%) | 1 (1.4%) | 0 (0%) |
| carcinosarcoma | 19 (20%) | 3 (19%) | 16 (22%) | 0 (0%) |
| chondrosarcoma | 1 (1.0%) | 0 (0%) | 1 (1.4%) | 0 (0%) |
| endometrial stromal sarcoma | 1 (1.0%) | 1 (6.3%) | 0 (0%) | 0 (0%) |
| fibrous sarcoma | 2 (2.1%) | 0 (0%) | 1 (1.4%) | 1 (17%) |
| gastrointestinal neuroendocrine carcinoma | 4 (4.2%) | 0 (0%) | 4 (5.4%) | 0 (0%) |
| gynecological clear cell carcinoma | 9 (9.4%) | 0 (0%) | 7 (9.5%) | 2 (33%) |
| leiomyosarcoma | 4 (4.2%) | 2 (13%) | 1 (1.4%) | 1 (17%) |
| liposarcoma | 1 (1.0%) | 0 (0%) | 1 (1.4%) | 0 (0%) |
| metaplastic breast carcinoma | 4 (4.2%) | 1 (6.3%) | 2 (2.7%) | 1 (17%) |
| neuroendocrine lung tumor | 2 (2.1%) | 0 (0%) | 2 (2.7%) | 0 (0%) |
| osteosarcoma | 1 (1.0%) | 0 (0%) | 1 (1.4%) | 0 (0%) |
| other thyroid cancer | 1 (1.0%) | 0 (0%) | 1 (1.4%) | 0 (0%) |
| ovarian serous carcinoma | 2 (2.1%) | 2 (13%) | 0 (0%) | 0 (0%) |
| pancreatic adenocarcinoma | 17 (18%) | 2 (13%) | 15 (20%) | 0 (0%) |
| pancreatic neuroendocrine tumor | 3 (3.1%) | 0 (0%) | 2 (2.7%) | 1 (17%) |
| peripheral nerve sheath tumor | 1 (1.0%) | 1 (6.3%) | 0 (0%) | 0 (0%) |
| prostate neuroendocrine carcinoma | 4 (4.2%) | 2 (13%) | 2 (2.7%) | 0 (0%) |
| salivary carcinoma | 4 (4.2%) | 1 (6.3%) | 3 (4.1%) | 0 (0%) |
| small bowel adenocarcinoma | 4 (4.2%) | 0 (0%) | 4 (5.4%) | 0 (0%) |
| synovial sarcoma | 1 (1.0%) | 0 (0%) | 1 (1.4%) | 0 (0%) |
| thymic squamous cell carcinoma | 1 (1.0%) | 0 (0%) | 1 (1.4%) | 0 (0%) |
| vascular sarcoma | 2 (2.1%) | 1 (6.3%) | 1 (1.4%) | 0 (0%) |
| well differentiated gastrointestinal neuroendocrine tumor | 2 (2.1%) | 0 (0%) | 2 (2.7%) | 0 (0%) |

| **Supplemental Table 2.** Histology for cancer types with representation in both IPS-H and -L | | | |
|---|---|---|---|
| **Cancer type** | **Overall**<br>**N = 52** | **IPS-High**<br>**N = 12** | **IPS-Low**<br>**N = 40** |
| carcinosarcoma | 19 (37%) | 3 (25%) | 16 (40%) |
| leiomyosarcoma | 3 (5.8%) | 2 (17%) | 1 (2.5%) |
| metaplastic breast carcinoma | 3 (5.8%) | 1 (8.3%) | 2 (5.0%) |
| pancreatic adenocarcinoma | 17 (33%) | 2 (17%) | 15 (38%) |
| prostate neuroendocrine carcinoma | 4 (7.7%) | 2 (17%) | 2 (5.0%) |
| salivary carcinoma | 4 (7.7%) | 1 (8.3%) | 3 (7.5%) |
| vascular sarcoma | 2 (3.8%) | 1 (8.3%) | 1 (2.5%) |
